# Neurogenic defects underlie functional bladder outflow tract obstruction associated with biallelic variants in *LRIG2*

**DOI:** 10.1101/2022.12.06.22283097

**Authors:** Celine Grenier, Filipa M. Lopes, Anna M Cueto-González, Eulàlia Rovira-Moreno, Romy Gander, Benjamin W Jarvis, Karen D. McCloskey, Alison M. Gurney, Glenda M. Beaman, William G. Newman, Adrian S. Woolf, Neil A. Roberts

## Abstract

Urinary tract malformations account for half of all children with kidney failure, and some have defined monogenic causes. One such disorder is urofacial, or Ochoa, syndrome (UFS), an autosomal recessive disease featuring a dyssynergic bladder with detrusor smooth muscle contracting against an undilated outflow tract. Incomplete voiding predisposes to urosepsis and kidney failure. Half of individuals with UFS carry biallelic variants in *HPSE2*, whereas some carry variants in *LRIG2* (*leucine rich repeats and immunoglobulin like domains 2*). We report one new kindred where the index case presented with fetal hydronephrosis and postnatally had urosepsis and functional bladder outlet obstruction. He had the grimace that, together with urinary tract disease, characterizes UFS. While *HPSE2* sequencing was normal, he carried a homozygous, predicted pathogenic, stop variant (c.1939C>T; p.Arg647*) in *LRIG2*. Hypothesizing that neurogenic defects underlie *LRIG2*-associated bladder dysfunction, we studied *Lrig2* homozygous mutant mice. Juveniles had enlarged bladders and *ex vivo* physiology experiments showed neurogenic defects in outflow tract relaxation. Mutants also displayed abnormal detrusor contractility. Moreover, there were nuanced differences in physiological defects between the sexes. The current case emphasizes that urinary tract disease in UFS begins before birth. Putting this family in the context of all reported urinary tract disease-associated *LRIG2* variants, the urinary and facial phenotype of UFS occurs with biallelic putative loss of function variants, but missense variants lead to bladder-limited disease without the grimace. Finally, our murine observations support the hypothesis that UFS is a genetic autonomic neuropathy of the bladder affecting outflow tract and bladder body function.

## INTRODUCTION

The mammalian urinary tract (UT) comprises the kidneys, ureters, bladder and urethra. Congenital UT malformations (UTMs) occur in 4 per 1000 human births (Bakker et al., 2018; Woolf, 2022) and UTMs account for approximately half of all children with severe kidney failure (Harambat et al., 2012). An estimated fifth of young adults with severe kidney failure were born with UTMs (Neild, 2009). Some individuals with a UTM have defined monogenic causes (Nigam et al., 2019; Jordan et al., 2022), and while most genetic research has focused on kidney malformations, monogenic causes of lower UTMs are now being discovered (Vivante et al., 2017; Woolf et al., 2019; Houweling et al., 2019; Mann et al., 2019; Beaman et al., 2022).

Urofacial, or Ochoa, syndrome (UFS), is an autosomal recessive condition causing lifelong ill health with potentially fatal complications (Ochoa 2004; Osorio et al., 2021). Affected females and males have dyssynergic bladders in which the detrusor smooth muscle (DSM) in the bladder body contracts against an outflow tract that fails to dilate fully. Thus, individuals with UFS experience frequent but incomplete urinary voiding. This functional bladder outflow obstruction leads to pooling of urine under high hydrostatic pressure within the bladder predisposing to vesicoureteric reflux, urosepsis, pyelonephritis and kidney failure (Ochoa 2004; Osorio et al., 2021). These UT features are accompanied by a characteristic grimace upon smiling, hence the term ‘urofacial’ (Ochoa 2004).

As reviewed (Newman and Woolf, 2018; Beaman et al., 2022), around half of genetically tested UFS individuals carry biallelic variants in *HPSE2* (OMIM UFS2 **#**236730), which encodes heparanase-2, a protein that binds heparan sulphate and also inhibits the endoglycosidase enzymatic activity of a related protein called heparanase (Levy-Adam et al., 2010; McKenzie et al., 2020).

Not all individuals with UFS have *HPSE2* variants. A small number of individuals with UFS have been reported to carry biallelic variants of *LRIG2* (OMIM USF2 **#**615112) (Stuart et al., 2013; Sinha et al., 2018). *LRIG2* codes for leucine rich repeats and immunoglobulin like domains 2, one of three mammalian LRIG proteins (Simion et al., 2014). Homozygous *Lrig2* or *Hpse2* mutant mice manifest aberrant urinary voiding (Stuart et al., 2015, Roberts et al., 2019), making them an appropriate model to study UFS in the laboratory.

We report here a new family with UFS, associated with a homozygous *LRIG2* variant, and place this in the context of all other UT disease-associated *LRIG2* variants reported to date. We hypothesized that peripheral neurogenic defects underlie LRIG2-associated bladder dysfunction. Accordingly, we studied *Lrig2* mutant mice with a battery of *ex vivo* physiology analyses. Our murine observations support the hypothesis that UFS is a genetic, autonomic neuropathy of the bladder with functional defects in both the outflow tract and DSM in the bladder body.

## MATERIALS AND METHODS

### Human Studies

The family described below was assessed at the Department of Clinical and Molecular Genetics and the Department of Pediatric Surgery, Pediatric Urology and Renal Transplant Unit, Hospital Vall D’Hebron, Barcelona, Spain. The parents, legal guardians, gave written informed consent, as approved by Vall d’Hebron Hospital Ethics Committee, for molecular genetic testing. The family also gave permission for their clinical story to be told.

### Mutant mice

*Lrig2* mice carrying a targeted deletion of exon 12 (Rondahl et al., 2013; Roberts et al., 2019) were maintained on a C57BL/6 background in the Biological Services Facility of The University of Manchester (Home Office Project Licence PP1688221). Homozygous wild-type (WT) and homozygous *Lrig2* (Mut) mice were generated by mating heterozygous parents. Lrig2 protein is present in normal mouse bladders but is not immunodetected in Mut bladders (Roberts et al., 2019).

### Ex vivo physiology on isolated bladder tissues

WT and Mut *Lrig2* mice were culled by cervical dislocation in accordance with Schedule 1 of the Animals (Scientific Procedures) Act 1986(ASPA) and with local AWERB approval (University of Manchester). Experiments were performed on bladder preparations from WT and Mut mice as described previously for *Hpse2* mutant mice (Manak et al., 2020). Briefly, the detrusor and outflow tracts were separated and mounted in myograph chambers (Danish Myo Technology, Hinnerup, Denmark) to measure isometric tension in physiological solution maintained at 37°C. Detailed protocols are described in the *Supplementary Methods*. Relaxation of the outflow tract is mediated by nitrergic nerves (Andersson & Persson 1993; Burnett et al., 1997; Manak et al., 2020). As incomplete dilatation of the bladder outflow tract is a key feature in people with UFS (Ochoa 2004), the ability of *Lrig2* Mut outflow tracts to relax in response to electrical field stimulation (EFS; 0.5 – 15 Hz, 80 V, 1 ms) or the directly acting nitric oxide (NO) donor, sodium nitroprusside (SNP), were compared with WT littermates. Relaxation was measured against a background of pre-constriction, which was evoked in males by the α-1 receptor agonist, phenylephrine (PE, 5 μM). Female outflow tracts contracted poorly to PE, confirming an earlier report (Alexandre et al., 2017). As female mouse urethras express transcripts encoding the AVP V1a receptor (Zeng et al., 2015), they were instead pre-contracted with arginine vasopressin (AVP). In separate experiments, outflow tracts or bladder bodies were stimulated with 50 mM KCl to directly depolarise the smooth muscle, which stimulates Ca^2+^ influx through voltage-gated Ca^2+^ channels to elicit contraction. Bladder bodies were subjected to EFS (0.5 – 25 Hz, 80 V, 1 ms) to induce frequency-dependent contractions mediated by acetylcholine-releasing parasympathetic nerves. Contraction was also evoked directly by the muscarinic agonist, carbachol (10 nM – 50 μM) and compared between WT and Mut mice.

### Histology

Histological evaluation of mouse bladder bodies was performed as previously described (Lopes et al., 2019; Roberts et al., 2019; Hindi et al., 2021; Ranjzad et al., 2020). We assessed anatomy by haematoxylin staining, collagen by picrosirius red (PSR) staining, and F4/80 macrophages and transforming growth factor β1 (TGFβ1) by immunostaining. Alignment of collagen fibrils was also assessed (Rezakhaniha et al., 2012). Detailed protocols are provided in the *Supplemental Methods*.

### Statistics

Details of data and statistical analysis are given in the *Supplemental Methods*.

## RESULTS

### Family history

The index case was a boy born to first cousin parents from Morocco. He has a grimace upon smiling, such that his eyes close and the corners of his mouth fail to elevate. The pregnancy leading to his birth was complicated by gestational diabetes mellitus, and bilateral hydronephrosis was detected on fetal ultrasound. At birth his weight and head circumference were within normal limits but increased body length was noted (55 cm, >99^th^ percentile) and attributed to gestational diabetes. His postnatal course featured urosepsis, and ultrasound revealed bilateral ureterohydronephrosis with a thickened bladder wall. Cystography revealed bilateral vesicoureteral reflux but the urethra appeared normal, excluding posterior urethral valves (**Figure 1A-C**). A clinical diagnosis of functional bladder outlet obstruction was made, and the bladder was drained by a percutaneous cystostomy device. Later in childhood, his psychomotor, language and learning development were normal, as were weight, height and head circumference. His plasma creatinine concentration was 0.41 mg/dl (36 μmol/l), within the normal range (0.24-0.73 mg/dl). *HPSE2* and *LRIG2* were sequenced (directed sequencing, Next Generation Sequencing). No variants were found in *HPSE2* but a homozygous variant (NM_014813.2): c.1939C>T (p.Arg647*), classified as pathogenic, according to American College of Medical Genetics guidelines (Richards et al., 2015), was identified in *LRIG2*. The stop codon was in exon 14 coding for an immunoglobulin-like domain (**Figure 1D**). The parents were healthy, heterozygous carriers. The index case has two older brothers who are clinically well: they have no urinary symptoms, lack a grimace upon smiling and have had normal abdominal ultrasound scans: their DNA was not available for testing. The variant in the index case is presented in the context of all other reported UT disease-associated variants in *LRIG2* in **Figure 1D** and **Table 1**.

**Figure 1.**
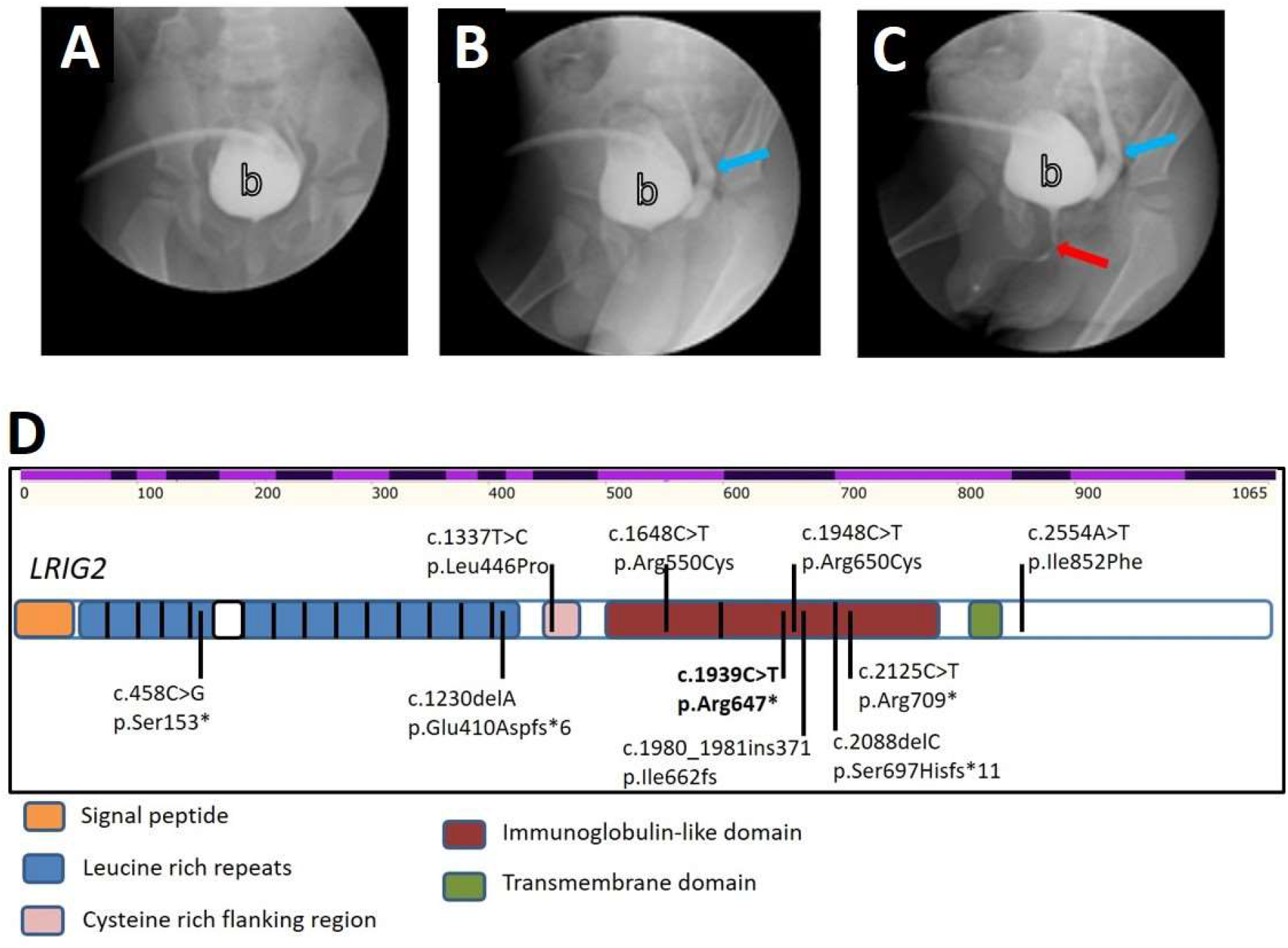
Identification of a novel pathogenic variant in *LRIG2* in a family affected by urofacial syndrome. **A-C**. Cystourethrogram sequential images as the bladder was filled *via* a suprapubic catheter, showing vesicoureteric reflux (blue arrow) and an anatomically patent urethra (blue arrow). **D**. Location of all identified *LRIG2* pathogenic variants associated with UT disease. The uppermost alternating light and dark purple row indicates positions of *LRIG2* exons and base position. Below this, the protein is depicted with its key domains indicated in different colours. Missense variants are shown above the protein sequence, while nonsense and frameshift variants are shown below the protein. The variant described in this paper is shown in bold and is located in the part of the gene coding for an immunoglobulin-like domain.

**Table 1.** UT disease-associated *LRIG2* variants reported to date. Frameshift and stop variants have been associated with full blown UFS, comprising UT disease and a grimace upon smiling. In contrast, missense variants are associated with UT disease without the grimace. All cases have homozygous variants apart from one compound heterozygous case, as indicated.

| Variant in *LRIG2* sequence | Predicted change in protein | Exon | Reference | Clinical features |
| --- | --- | --- | --- | --- |
| c.458C>G | Stop variant<br>p.Ser153* | 4 | Sinha et al 2018 | UFS (grimace and urinary tract disease) |
| c.1230delA | Frameshift variant<br>p.Glu410Aspfs*6 | 10 | Stuart et al 2013 | UFS (grimace and urinary tract disease) |
| c.1337T>C | Missense variant<br>p.Leu446Pro | 12 | Roberts et al 2019 | Urinary tract disease (without UFS grimace) |
| c.1648C>T | Missense variant<br>p.Arg550Cys | 13 | Stuart et al 2013 | Urinary tract disease (without UFS grimace) |
| c.1939C>T | Stop variant<br>p.Arg647* | 14 | This study | UFS (grimace and urinary tract disease) |
| c.1948C>T | Missense variant<br>p.Arg650Cys | 14 | Roberts et al 2019 | Urinary tract disease (without UFS grimace) |
| c.1980_1981ins371<br>GenBank JX891452<br>and<br>c.2088delC | Frameshift variant<br>p.Ile662fs<br>and<br>Stop variant<br>p.<br>Ser697Hisfs*11 | 14 | Stuart et al 2013 | UFS (grimace and urinary tract disease) |
| c.2125C>T | Stop variant<br>p.Arg709* | 15 | Stuart et al 2013 | UFS (grimace and urinary tract disease) |
| c.2125C>T | Stop variant<br>p.Arg709* | 15 | Fadda et al 2016 | Urinary tract disease (without classic UFS grimace but with other facial dysmorphology) |
| c.2554A>T | Missense variant<br>p.Ile852Phe | 16 | Stuart et al 2013 | Urinary tract disease (without UFS grimace) |

### General characteristics of Lrig2 mutant mice and their bladders

To learn more about the underlying pathophysiology of *LRIG2* UFS, we studied *Lrig2* homozygous Mut mice. Given potential differences between the sexes, male and female mice were analysed separately. Juvenile males were studied between 14 and 19 days after birth, with no significant difference in ages between WT and Mut groups **(Figure S1A)**. Juvenile females were studied between 14 and 20 days, with no significant difference in ages between WT and Mut groups (**Figure S1B**). Male Mut mice weighed significantly less than their WT littermates (**Figure S1C**), and female Mut mice weighed significantly less than WT (**Figure S1D**). Inspection on autopsy indicated that, in each sex, Mut bladders were more prominent than WT bladders, with Mut bladders typically distended with urine (**Figure 2A**), as previously reported (Roberts et al., 2019). There was no significant difference between the two genotypes in the weights of male bladders (i.e. body plus outflow tract) drained of urine (**Figure S1E**) but the bladder/body weight ratio (**Figure 2B**) was significantly higher in male mutants. The weight of female bladders drained of urine was greater in the Mut *versus* WTs (**Figure S1F**). Female bladder/body weight ratio (**Figure 2B**) was also higher in Mut mice.

**Figure 2.**
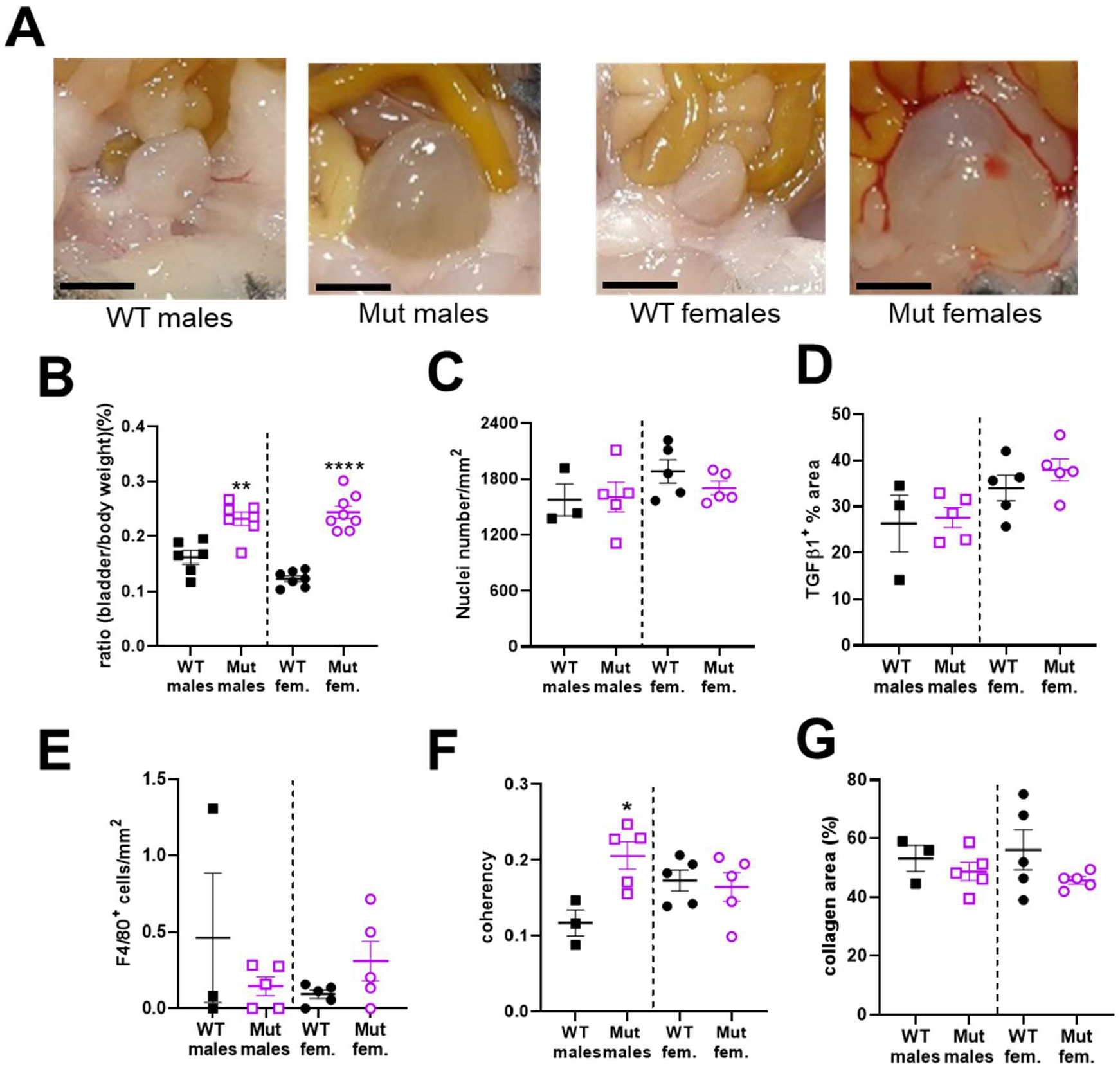
WT and *Lrig2* Mut bladders. **A**. Representative bladders at autopsy. Note the Mut bladders were distended with urine. Scale bars are 2 mm. **B**. Bladder/body weight ratio was higher in Mut *versus* WT males (WT n=6, Mut n=7, p=0.0018), and also in Mut *versus* WT female littermates (WT n=6, Mut n=8, p<0.0001). **C**. Nuclei number/area in the detrusor was similar between WT and mutant males (WT n=3, Mut n=5, p=0.905), and between WT and mutant females (WT n=5, Mut n=5, p=0.2506). **D**. Percentage area positive for TGFβ1 staining in the detrusor was not different between male WT and mutant mice (WT n=3, Mut n=5, p=0.819). It was also similar between female WT and mutant (WT n=5, Mut n=5, p=0.315). **E**. Cell numbers positive for the macrophage marker F4/80 in the detrusor were similar between male WT and mutants (WT n=3, Mut n=5, p=0.3569) and between female WT and mutants (WT n=5, Mut n=5, p=0.140). **F**. Coherence of collagen fibrils in the detrusor was higher in male Mut compared with WT (WT n=3, Mut n=5, p=0.0204) but it was similar between female WT and mutants (WT n=5, Mut n=5, p=0.7256). **G**. The percentage area positive for collagen staining in the detrusor was similar between male WT and mutants (WT n=3, Mut n=5, p=0.434) but was similar between female WT and mutants (WT n=5, Mut n=5, p=0.168). In this and in subsequent figures, WT males are represented by black filled squares, Mut males by purple open squares, WT females by black filled circles and Mut females with purple open circles. Results are expressed as mean±SEM. *p<0.05, ** p <0.01, ****p<0.0001 are the significant differences between Mut and WT of the same sex.

### Bladder histology

Given the enlargement of Mut bladders, we investigated histological differences between Mut and WT bladders. Bladder domes sectioned and stained with haematoxylin had similar general appearances (**Figure S2A**). The numbers of detrusor nuclei per area did not differ between the genotypes in males (**Figure 2C**) or in females (**Figure 2C**), suggesting a lack of significant cell hypertrophy. To determine whether inflammation or fibrosis might be present, TGFβ1, macrophages and collagen were assessed. The percentage of detrusor area immunostaining for TGFβ1 (**Figure S2B**) was similar (**Figures 2D**) between the genotypes in males and females. The numbers of macrophages in the detrusor (**Figure S2C** and **Figure 2E**) was also similar in the two genotypes. From PSR staining and bright field imaging (**Figure S3A)**, the percentage area positive for collagen in detrusor was similar between the genotypes (**Figure 2G**) for both males and females. PSR was also imaged under polarised light to observe the orientation, or ‘coherency’ (Finlay et al., 1995) of collagen fibrils (**Figure S3B and Figure 2F**). In male detrusor, fibril coherency was significantly higher in mutants but no difference was found between genotypes in females.

### Physiology of isolated bladder outflow tracts

Having established that the Mut bladders were not overtly fibrotic or inflamed, we proceeded to undertake *ex vivo* physiology experiments on bladder outflow tracts using myography. In males, no significant difference in contraction amplitude to 50 mM KCl between Mut and WT tissues was observed (**Figure 3A**). As shown in **Figure S4**, PE evoked contraction of male Mut outflows with similar potency and efficacy to WT preparations. The relaxation response to the NO donor SNP (**Figure 3B**) was also similar in Mut and WT outflow tracts. In contrast, EFS evoked frequency-dependent relaxation of PE-contracted outflows that were significantly smaller across all frequencies in Mut tissues compared to WT (**Figure 3C** and **3D**). Outflow tracts from females with Mut and WT genotypes had similar amplitude contractions in response to 50 mM KCl (**Figure 3E**) and relaxed with similar responses to SNP (**Figure 3F**). A striking difference was observed, however, in their responses to EFS (**Figure 3G and H**). While female WT outflows pre-contracted with AVP displayed frequency-dependent relaxation in response to EFS, Mut outflows showed frequency-dependent contraction. Contractions at all frequencies was blocked by pre-incubating Mut outflows with tetrodotoxin, which prevents neurotransmission in the tissue by blocking Na channels.

**Figure 3.**
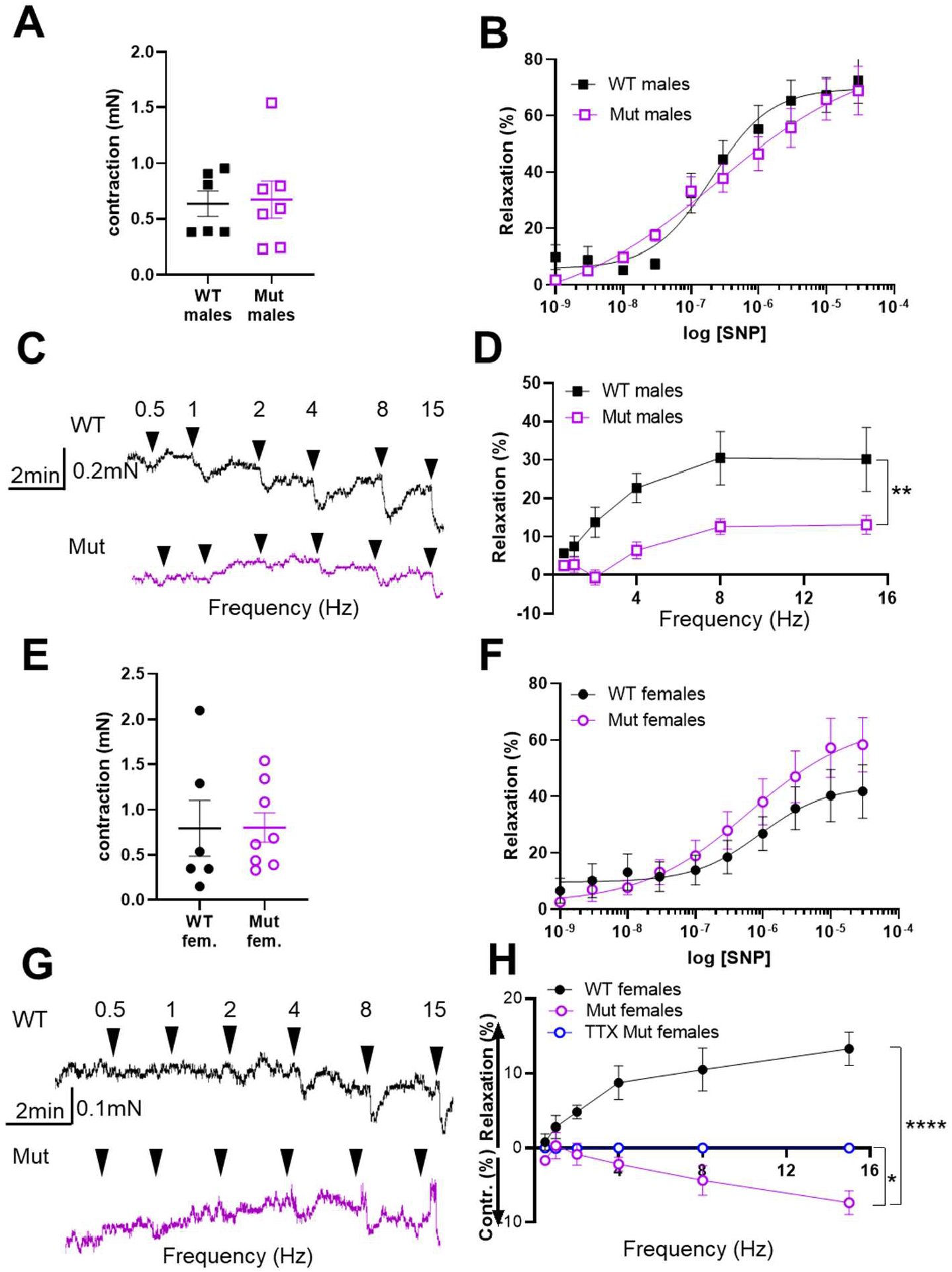
Dysfunctional outflow physiology in *Lrig2* mutant mice. **A**. Amplitudes of contractions evoked by 50 mM KCl in male WT (n=6) and Mut (n=7) outflow tracts. **B**. Outflow relaxations in response to cumulatively increased concentrations of SNP in WT (n=6) and mutant (n=6) male outflow tracts. Curves show best fits to the Hill equation with EC_50_ = 0.20 μM (control) and 0.24 μM (Mut) and E_max_ = 81.7 % (control) and 80.6 % (Mut). **C**. Representative traces of WT and mutant male outflows contracted with 5 μM PE then stimulated by EFS at the frequencies indicated. **D**. Mean ± SEM relaxations evoked by EFS, plotted as a function of frequency in male WT (n=5) and Mut (n=7) outflows. ** p=0.005 **E**. Amplitudes of contraction evoked by 50 mM KCl in female outflows (WT n=6, Mut n=8). **F**. Female outflow relaxations in response to cumulatively increased concentrations of SNP in WT (n=6) and mutant (n=8) outflow tracts. Curves show best fits to the Hill equation with EC_50_= 0.91 μM (control) and 0.61 μM (Mut) and E_max_ = 43.7 % (control) and 66.5 % (Mut). **G**. Representative traces of WT and mutant female outflows contracted with 10 nM AVP then stimulated by EFS at the frequencies indicated. **H**. Mean responses to EFS as a function of frequency in female outflows (WT n=6, Mut n=8). Responses of Mut outflows tested in the presence of TTX (N=4) are also shown. Results are expressed as mean ± SEM. * p= 0.02 Mut *vs* TTX, **** p<0.0001 WT compared with Mut.

### Physiology of ex vivo bladder body preparations

Rings of bladder bodies from Mut and WT male mice contracted equally in response to the application of 50 mM KCl (**Figure 4A**). In contrast, bladders from Mut male mice were more sensitive to carbachol than those from WT males (**Figure 4B**), with a significantly higher dose-response curve to carbachol. Bladder rings from male mice contracted in response to EFS, the contraction increasing in amplitude with stimulation frequency (**Figure 4C**). Despite their increased responsiveness to carbachol, responses to EFS were smaller in bladders from Mut mice compared with WT mice (**Figure 4D**). Bladder rings from female Mut mice contracted less forcefully to 50 mM KCl than rings from WT females (**Figure 4E**). Rings of Mut female bladder also contracted less to carbachol than WT female bladder, in Mut mice (**Figure 4F**), with a significantly lower dose-response curve to carbachol. EFS evoked frequency dependent contraction in bladder rings from female WT and Mut mice, but the responses of Mut bladders were much weaker (**Figure 4G and 4H**).

**Figure 4.**
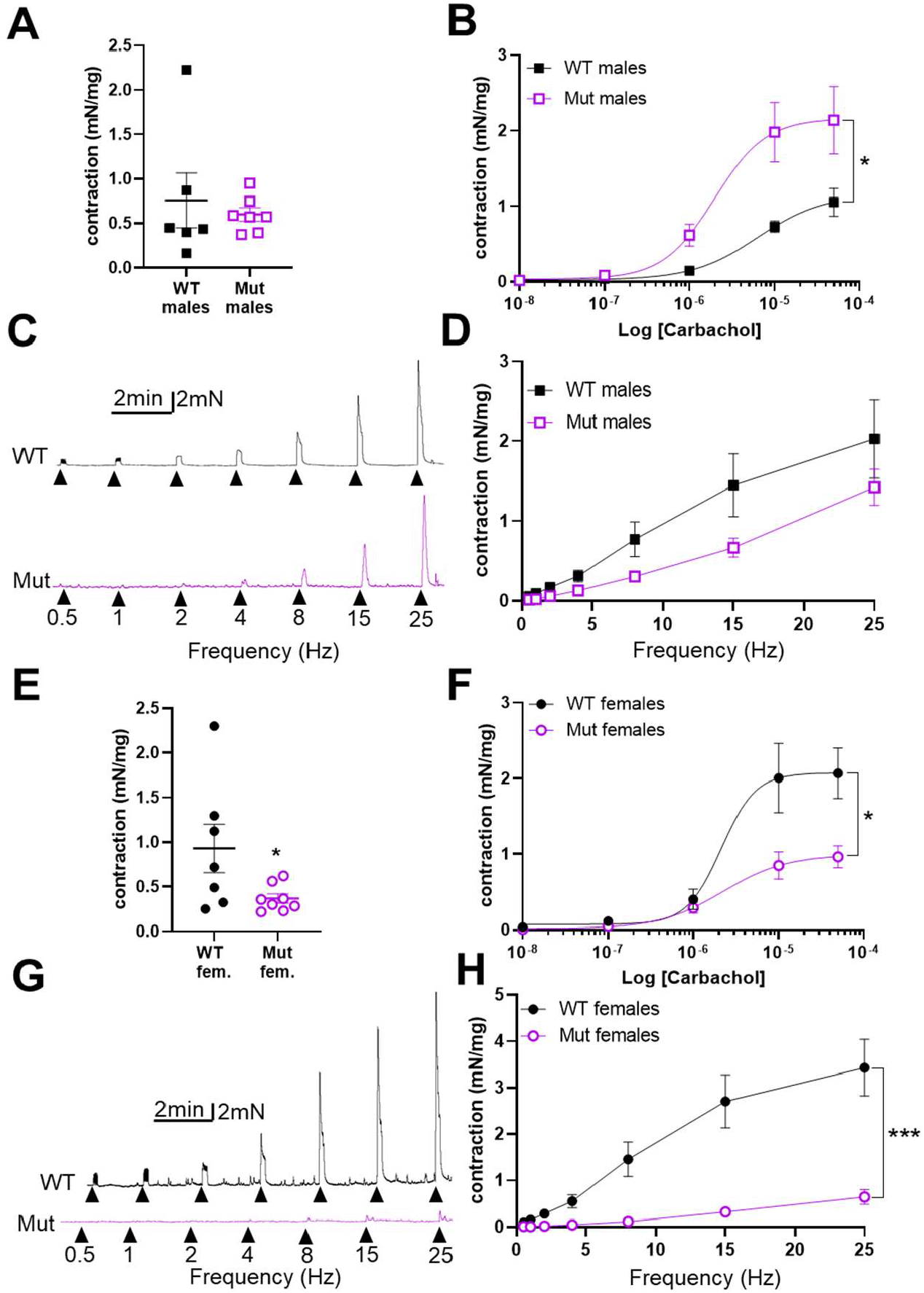
Dysfunctional bladder body physiology in *Lrig2* mutant mice. **A**. Amplitudes of contractions evoked by 50 mM KCl in bladder rings from WT (n=6) and Mut (n=7) male mice. **B**. Contraction of bladder rings from WT (n=5) and Mut (n=7) male mice in response to cumulative application of 10 nM – 50 μM carbachol, plotted as a function of carbachol concentration. Curves are the best fits of the Hill equation with EC_50_ = 6.37 μM and E_max_ = 1.15 mN/mg in WT mice compared with EC_50_ = 1.97 μM and E_max_ = 2.16 mN/mg in Mut mice. * p = 0.04 comparing WT and Mut by 2-way ANOVA **C**. Representative traces of contraction evoked in male WT and Mut bladder rings in response to EFS at the frequencies indicated. **D**. Mean amplitudes of contraction evoked by EFS in bladders from male WT (n=5) and Mut (n=7) mice plotted as a function of frequency. **E**. Amplitudes of contractions evoked by 50 mM KCl in bladder rings from WT (n=7) and Mut (n=8) female mice. **F**. Contraction of bladder rings from WT (n=7) and Mut (n=8) female mice in response to cumulative application of 10 nM – 50 μM carbachol, plotted as a function of carbachol concentration. Curves are the best fits of the Hill equation with EC_50_ = 2.13 μM and E_max_ = 2.07 mN/mg in WT mice compared with EC_50_ = 2.26 μM and E_max_ = 0.99 mN/mg in Mut mice. * p = 0.02 comparing WT with Mut by 2-way ANOVA. **G**. Representative traces of contraction evoked in female WT and Mut bladder rings in response to EFS at the frequencies indicated. **H**. Mean amplitudes of contraction evoked by EFS in bladders from female WT (n=5) and Mut (n=7) mice plotted as a function of frequency. *** p = 0.0006 comparing WT and Mut by 2-way ANOVA. Results are expressed as mean ± SEM.

## DISCUSSION

This study advances our understanding of the genetics and pathophysiology of congenital bladder outflow obstruction, combining insights from human disease and a mouse model.

### Insights into LRIG2 associated human bladder disease

We describe a variant in *LRIG2* in a hitherto unreported family with UFS. The variant in exon 14 results in a stop codon in the second immunoglobulin-like domain of the extracellular part of the protein. Putting this new family in the context of all reported UT-disease associated *LRIG2* variants, as depicted in Figure 1D and Table 1, it is evident that biallelic stop or frame shift variants, consistent with a putative loss of function mechanism, can be associated with a complete UFS phenotype, combining the characteristic grimace with UT disease, as reported in this and other (Stuart et al., 2013, Sinha et al., 2018) studies. This relationship is not exact because one study reported a stop variant with marked UT disease but without the typical inverted smile (Fadda et al., 2016); that individual, however, did have an unusual smile and other facial dysmorphology. In contrast, individuals with biallelic missense variants (Stuart et al., 2013; Roberts et al., 2019) uniformly lack the UFS facial grimace but have severe functional bladder outlet obstruction. Further research is needed to explain these genotype/phenotype relationships. However, the unified message to clinicians is to consider *LRIG2* and *HPSE2* variants as a cause of congenital bladder obstruction whether a UFS grimace is present or not. The patient we describe in the current report was exposed to gestational diabetes mellitus. Human clinical studies as well as animal experiments suggest that maternal diabetes mellitus can perturb UT development (Dart et al., 2015; Groen In’t Woud et al., 2016; Hokke et al., 2013). The phenotypes associated with maternal diabetes, however, are kidney hypoplasia or agenesis rather than functional bladder outflow obstruction. Therefore, the UT phenotype in the current case is highly unlikely to have been modified by maternal diabetes.

### Initial evidence that a peripheral neuropathy underlies UFS

Individuals with UFS usually present with dysfunctional urinary voiding and urosepsis in early childhood (Ochoa 2004; Osorio et al., 2021), and it is also recognized that some such patients have large bladders visualized before birth (Newman and Woolf, 2018). Indeed, the individual described in this study had all of these features. Key questions are, therefore, to understand the biological and physiological roles of *LRIG2* and *HPSE2* in healthy bladders, and how these processes go wrong in the presence of germline variants to generate the clinical phenotypes. LRIG2 and heparanase-2 proteins have been detected in nerves within the late first trimester in human fetal bladder (Stuart et al., 2013), and in their migrating neural crest precursors in earlier human embryos (Beaman et al., 2022). Moreover, both proteins are present in mouse pelvic ganglia (Stuart et al., 2015; Roberts et al., 2019) which send postganglionic autonomic neurons into the lower UT (Keast et al., 2015; Roberts et al., 2019). Homozygous *Lrig2* or *Hpse2* mutant mice both have abnormally patterned bladder nerves (Roberts et al., 2019). Full length LRIG2 is a plasma membrane protein but a soluble cleaved form has also been described (Kim et al., 2021). LRIG2 enhances tumour growth in diverse tissues, including brain and skin, a property conferred in part by modulating growth factor signaling (Rondahl et al., 2013; Dong et al., 2020; Hoesl et al., 2019). Mouse studies have also implicated LRIG2 in axon guidance during central nervous system development and regeneration (van Erp et al., 2015). Collectively, these observations led us to predict that LRIG2 has important roles in the physiology of bladder components implicated in normal voiding, with defects leading to functional bladder outflow obstruction.

### Lrig2 mutant mice have neurogenic defects in bladder outflow tract relaxation

For optimal voiding to occur, the outflow tract must relax when the bladder body contracts. NO released from nitrergic nerves is considered the primary mechanism of outflow tract relaxation during micturition in mammals (Andersson et al., 1993). In male *Lrig2* mutant mice, we observed reduced nitrergic-nerve mediated outflow relaxation. The outflow maintained its responsiveness to SNP, implying that the loss of nitrergic relaxation in the mutant outflow was due to less NO reaching the muscle rather than an altered NO action. This accords with a report that *Lrig2* mutant mouse outflow tracts appears to contain fewer nitrergic nerves than WT controls (Roberts et al., 2019). Male *Lrig2* mutant outflows were otherwise ‘physiologically normal’. Contractions induced by KCl or the α1 receptor agonist PE were preserved in mutant outflow tracts, again indicating that non-neurogenic contraction of the outflow SM is normal in *Lrig2* Mut mice. Cautiously extrapolating to humans, male UFS patients might be predicted to have a functional bladder outflow obstruction of neurogenic origin, specifically with diminished nitregic nerve function.

Female *Lrig2* mutant mouse outflow tracts differed in detail from the male mutants, showing a more dramatic loss of nitrergic relaxation, yet the functional effect would be the same i.e. bladder outflow obstruction. Indeed, reviewing the collective but limited literature (the current study; Stuart et al., 2013; Sinha et al., 2018), males and females with UFS and *LRIG2* variants have UT disease of apparently similar severity. We used a modified technique to investigate neurogenic function in female mice, because WT female outflows did not contract to α_1_-receptor agonists, possibly due to the absence of sympathetic nerves, neurotransmitter release or receptors. Female outflow tracts were instead contracted with AVP to enable relaxation to be studied, as previously described (Zeng et al., 2015). In that context, EFS evoked relaxation responses that were frequency-dependent and TTX-sensitive in female WT outflow tracts, as seen in males (Manak et al, 2020). Strikingly, EFS evoked further contraction, rather than relaxation, of *Lrig2* mutant female outflows precontracted with AVP. These contractions were also tetrodotoxin-sensitive, indicating a neuronal origin but we were unable to identify the neurotransmitter involved. Translating this finding to humans is subject to caveats, not least because female neuro-urological anatomy varies between species. Although the outflow tracts of female mice lack a sympathetic contractile response, this may not be typical of larger mammals. For example, α_1_-receptor stimulation induced muscle contraction in urethral preparations from female pigs (Bridgewater et al., 1995) and humans (Taki et al., 1999). Moreover, similar levels of α_1_-receptor transcripts were detected in female and male outflow tracts from patients with invasive bladder cancer (Nasu et al., 2009). Pigs may therefore be a better model than mice for human bladder physiology.These considerations point to further work being needed to test the physiology of bladder outflow tracts in healthy male and female humans, and in individuals with UFS. A working hypothesis is that in both males and females with UFS there is defective urethral dilation, but with differing underlying pathophysiology. If this is correct, then different types of drug may be optimal to help bladder emptying in males or females with UFS.

### Lrig2 mice also have abnormal detrusor contractility

Our investigation of the bladder body of *Lrig2* mutant mice also revealed a strong neurogenic phenotype. Frequency-dependent contractions evoked by EFS were smaller in the mutant bladder preparations, with the difference between WT and mutant more pronounced in females than males. These findings are broadly consistent with a previous study of juvenile male *Hpse2* mutant mice (Manak et al., 2019). On the other hand, effects of the *Lrig2* mutation on muscle contraction evoked directly by membrane depolarization or receptor stimulation diverged between the sexes. The male *Lrig2* mutant bladder generated a normal level of contraction in response to depolarization with KCl but was hyper-responsive to muscarinic receptor-mediated stimulation. In female mice, contractile responses to both KCl and muscarinic receptor stimulation were suppressed in the *Lrig*2 mutant bladder body. The muscarinic M3 receptor is the predominant mediator of muscarinic stimulation in mouse and human bladder bodies (Chopin et al., 2002, Chess-Williams et al., 2001; Fetscher et al., 2002) and is activated by acetylcholine released from parasympathetic nerves. This activates G-proteins and their downstream signaling cascades to induce bladder contraction. The hypersensitivity to carbachol in male Mut mice could be due to upregulation of this pathway, rather than a general increase in contractility, because it was not seen with KCl-induced contraction. It could be a primary mechanism of UFS pathology, or a secondary response to the loss of parasympathetic input. Further work is needed to assess these possibilities. Whatever the mechanism, it was not seen in females, which rather showed a general loss of detrusor power, analogous to bladder decompensation reported in long-term diabetes mellitus (reviewed in Hindi et al., 2020).

The apparently more severe *ex-vivo* physiological defects in female *Lrig2* mutant bladder bodies, compared with males, correlates with the more marked change in bladder weight found in female mutants. While in both sexes the bladder to whole body weight ratios were increased compared to WT mice, only the female mutants had an absolute increase in bladder weight compared with the WT controls. This difference may be a primary feature of the disease or, perhaps more likely, it reflects a more marked growth response to outflow obstruction in females than was present in males. In general, differences between males and females in their physiological responses could be a direct consequence of the mutation in *Lrig*2, or they could occur secondary to outflow obstruction. The current study could not address this question, but we note that there was no evidence of a fibrotic or inflammatory process in the DSM of either sex on histology. The only difference found between mutants and controls was an increased collagen coherence in male mutant DSM, which may reflect higher hydrostatic pressures in these bladders, by analogy with collagen coherence studies in (non-mutant) arteries (Finlay et al., 1995).

## CONCLUSIONS

The study highlights several notable clinical, genetic and pathobiological points. The current case constitutes further evidence that biallelic *LRIG2* variants can cause UFS. It also emphasises that UT disease in UFS begins before birth. Putting this family in the context of all reported UT disease-associated *LRIG2* variants reported, full-blown UFS occurs with loss of function variants, but missense variants lead to bladder-limited disease without the grimace. For the clinician, it is important to consider *LRIG2* and *HPSE2* variants as a cause of congenital bladder obstruction whether a UFS grimace is present or not. Moreover, our murine observations support the idea that UFS is a genetic autonomic neuropathy of the bladder affecting functionality of the outflow tract and the bladder body. The nuanced sex differences in abnormal *ex-vivo* physiology in *Lrig2* mutant mice may suggest that drug treatments could be tailored for males or females with UFS and should prompt further studies of possible sex differences in bladder outflow and body physiology in humans.

## Data Availability

All data produced in the present work are contained in the manuscript

## ACKNOWLEDGEMENTS

We acknowledge receiving research funding from: Medical Research Council (project grant MR/L002744/1 to ASW and WGN; project grant MR/T016809/1 to ASW, NAR, and FML; and Doctoral Training Programme to BWJ); Kidney Research United Kingdom (project grant Paed_RP/002/20190925 to WGN, GMB, and ASW; and Paed_RP/005/20190925 to NAR and ASW); Newlife Foundation (project grants 15-15/03 and 15-16/06 to WGN and ASW); the Manchester NIHR Biomedical Research Centre (IS-BRC-1215-20007 to WGN); and Kidneys for Life (start-up grant 2018 to NAR). We thank the European Reference Network on Rare Congenital Malformations and Rare Intellectual Disability ERN-ITHACA (EU Framework Partnership ID: 3HP-HP-FPA ERN-01-2016/739516) for their support. We thank the Manchester NIHR Biomedical Research Centre Rare Diseases theme, and the Manchester Rare Condition Centre for support.

