## Supplementary for "Neurogenic defects underlie functional bladder outflow tract obstruction associated with biallelic variants in *LRIG2*"

**Supplementary Methods**

**Ex-vivo physiology**

Mice were culled by cervical dislocation in accordance with Schedule 1 of the Animal (Scientific Procedures) Act, 1986. The bladder and its outflow tract, containing the urethra, were dissected as a unit and placed in cold physiological salt solution (PSS; 122 mM NaCl, 5 mM KCl, 10 mM N-2-hydroxyethylpiperazine-N'-2-ethanesulfonic acid (HEPES), 0.5 mM KH_2_PO_4_,1 mM MgCl_2_, 5 mM D-glucose, and 1.8mM CaCl_2_) and adjusted to pH 7.3 with NaOH. Connective tissues were removed from outflow tracts, which were then with a cut below the bladder neck and above the striated muscle of the external sphincter. Outflow tracts were mounted between the pins of a Myograph chamber (Danish Myo Technology, Hinnerup, Denmark) containing PSS maintained at 37°C and bubbled with air. The pins were separated by 50 µm and the outflow tracts equilibrated for 20 min, during which time a spontaneous and sustained contraction developed. The optimal tension to produce the largest contractile response from the tissue was determined as follows. Each tissue was stimulated with either 5 µM PE (males) or 80 mM KCl (females) until the change in tension reached a plateau, when it was washed. Between each stimulation the pins were separated by a further 5 µm until two successive contractions had the same amplitude. After washing and 10 min rest, 50 mM KCl was added to test the receptor-independent ability of outflow tracts to contract. They were then washed and pre-contracted with 5 μM PE (males) or 10 nM AVP (females). Once the contraction reached a plateau, EFS (1 ms 80 V pulses at 5 - 15 Hz for 10 s) was applied using a Grass SD9 stimulator (Grass Instruments, USA) to activate neurogenic relaxation. An interval of 2 min was allowed for recovery between each stimulation. In some experiments, the Na channel blocker tetrodotoxin (TTX, 1 nM) was applied before delivering EFS to inhibit firing of neuronal action potentials and prevent neurotransmitter release.

To assess the responsiveness of male outflow tract to phenylephrine (PE), it was applied cumulatively at 10 nM to 200 µM and concentration-response curves constructed from the response amplitudes. To assess the effect of sodium nitroprusside (SNP), it was applied cumulatively to precontracted outflow tracts at concentrations of 1 nM to 10 µM. Stock solutions of SNP were stored on ice in the dark and diluted into PSS immediately before use. Precontraction was generated by 5 µM PE in male outflow tracts or by 10 nM AVP in female tissues.

Bladder bodies, isolated from outflow tracts, were weighed before a 1.5 - 2.0 mm wide ring was cut and mounted between pins in a myograph chamber. A basal tension of 2 mN was applied and the bladder ring left to equilibrate for 20 min. To confirm the physiological viability of the tissue, a single application of 10 µM carbachol was applied, contraction recorded, and the tissue then washed. After 10 min recovery, receptor-independent contraction was induced by adding 50 mM KCl. Once contraction reached a peak, the tissue was washed and tension allowed to return to baseline before increasing frequencies of EFS (0.5 Hz – 25 Hz) were applied, with a 2 min gap between each stimulation. After a 10 min period for recovery, carbachol (10 nM - 50 µM) was applied cumulatively, allowing any tension generated to reach a plateau before the next addition. Contraction amplitudes for individual specimens were normalised to the bladder ring weights and plotted as a function of carbachol concentration.

The maximum contraction or relaxation response (E_max_) and the concentration evoking 50% of the maximum response (EC_50_) were estimated by fitting concentration-response plots to the Hill equation:

$$E=\frac{E_{max}-E_{min}}{1+\left( \frac{EC_{50}}{\left[ X \right]} \right)^{n}}$$

where E is the amplitude of the effect (contraction or relaxation), E_min_ is the minimum effect or response, X the concentration of drug and *n* the Hill slope, quantifying the slope of the steepest part of the concentration-response relationship.

***Collagen staining***

Domes from the bladders that were used for physiology experiments were fixed with 4% PFA, and embedded in paraffin, as previously described (Roberts et al., 2019). Sections (5 µm thickness) were cut using a microtome (Leica). After dewaxing and rehydration, sections were stained for 60 minutes with picrosirius red (PSR; Abcam), then dehydrated, cleared and mounted using Entellan mounting medium. Images were collected on an Olympus BX63 upright microscope using a 10x / 0.40 UPlanSApo objective and captured and white-balanced using a DP80 camera (Olympus) in RGB mode through CellSens Dimension v1.16 (Olympus). Images were then processed and analysed using Fiji ImageJ (Hindi et al., 2021) PSR was also visualised under cross-polarised light, to detect birefringent collagen, and alignment of the collagen fibrils (Rezakhaniha et al., 2012) using an Image J plugin OrientationJ, as described (Hindi et al., 2021).

***F4/80 macrophage immunostaining***

After dewaxing and rehydration, sections were treated with 1% hydrogen peroxidase for 20 minutes to block endogenous peroxidase activity. Antigen retrieval was performed in boiling citrate buffer. Sections were then incubated with primary antibody (rabbit anti F4/80 [SP115] ab111101 Abcam), at 1:100 dilution in PBS, 0.1% Triton X-100, 3% goat serum, in humid chambers overnight at 4°C. After a series of washes, a biotinylated goat anti-rabbit secondary antibody (ab6720, Abcam) diluted 1:200 in PBS was added for 1 hour at room temperature. After washing, sections were incubated with Vectastain ABC Reagents (Vector Laboratories) for 30 minutes. Sections were then developed in DAB (Vector Laboratories) for 2 ½ minutes to detect peroxidase activity and counterstained with haematoxylin for 30 seconds. The slides were then dehydrated, cleared and mounted using Entellan mounting medium (Sigma Aldrich). Images were collected using a 4x / UPlanFL N and a 40x / UApo/340 C1 objective, and captured and white-balanced using a DP80 camera (Olympus) in RGB mode through CellSens Dimension v1.16 (Olympus). Images were then processed and analysed using Fiji ImageJ. The images were scaled, and the detrusor area delimited, and F4/80 positive cells quantified and normalised per mm^2^.

***TGF****β****1 Immunostaining***

Slides were prepared as for F4/80 staining then stained using a Mouse-on-Mouse (M.O.M.) block kit (Vector Laboratories). Sections incubated in M.O.M Mouse IgG blocking reagent for 1 hour at room temperature. Sections were incubated overnight in primary antibody (mouse anti-TGFβ1, sc-130648 SantaCruz) 1:400 in M.O.M diluent, and incubated overnight at 4°C in a humid chamber. Sections were then washed and incubated for 10 minutes with M.O.M. Biotinylated anti-mouse reagent diluted to 1:200 in M.O.M diluent. After washing, the sections were incubated with Vectastain ABC Reagents (Vector Laboratories) for 30 minutes. They were then developed in DAB (Vector Laboratories) for 2 min, 30 s minutes to detect peroxidase activity, and counterstained for 30 seconds with haematoxylin. Slides were then dehydrated, cleared and mounted using Entellan mounting medium (Sigma Aldrich). Images were collected using a 20x UApo/340 objective and captured and white-balanced using a DP80 camera (Olympus) in RGB mode through CellSens Dimension v1.16 (Olympus). Images were processed and analysed using Fiji ImageJ. The images were scaled, and the detrusor area delineated?. Colour deconvolution was used to separate the brown colour of the positive signal. The resultant image was then thresholded and the percentage of area stained determined.

***Statistical analyses***

Data organised in column are expressed as mean±SEM and plotted and analysed using the GraphPad Prism 8 software. Normality of data distributions were assessed using Shapiro-Wilk test. If the values in two sets of data passed this test and they had the same variance, a Student t-test was used to compare them. If the variance was different, a t-test with Welch’s correction was used. If data did not pass the normality test, a non-parametric Mann-Whitney test was used. Groups of data with repeated measurements (e.g. at different stimulation frequencies or concentration) were compared using 2-way ANOVA with repeated measures. An F-test was used to assess the likelihood that independent data sets forming concentration-response relationships were adequately fit by a single curve.

**Supplementary Figures**

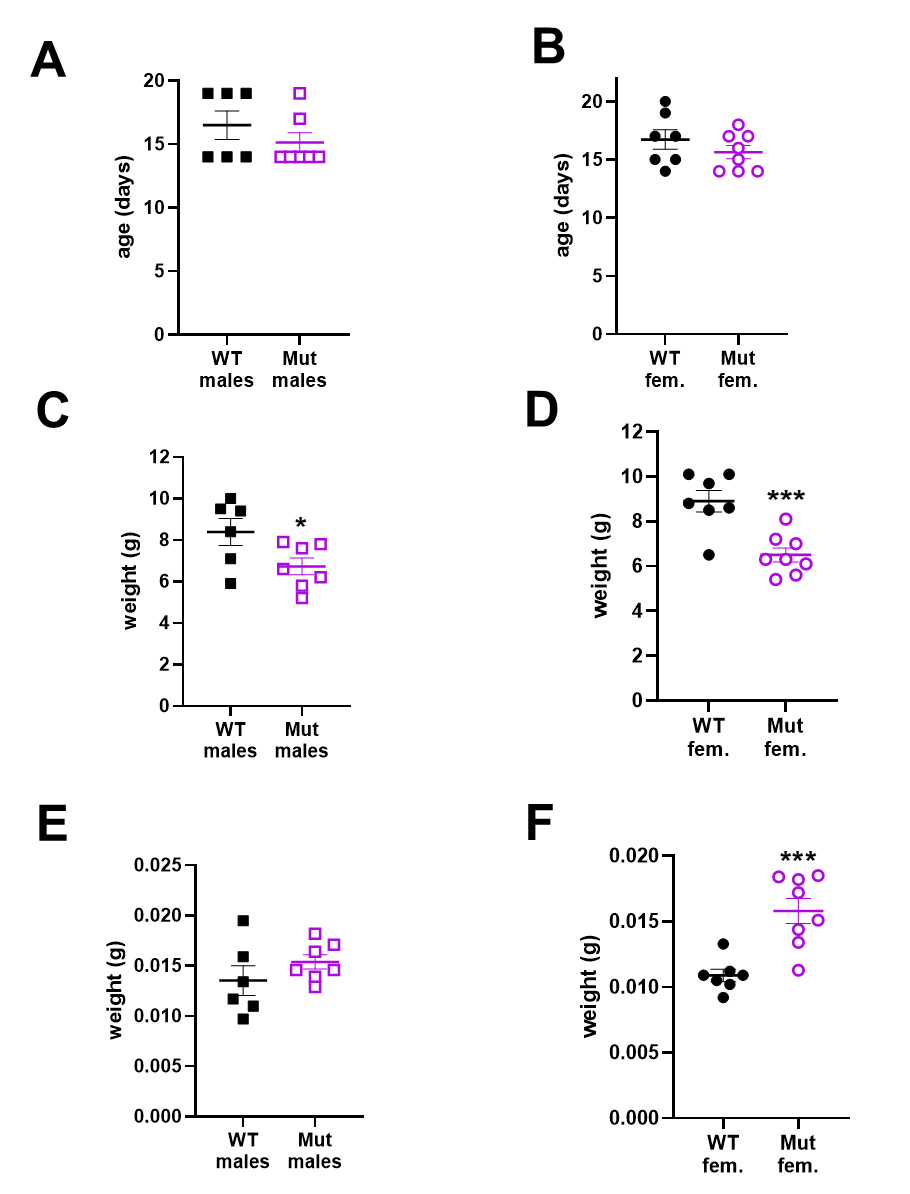

**Figure S1. Mouse ages and bladder weights.** **A.** WT and *Lrig2* Mut males mice were studied at similar ages (WT n=6, Mut n=7, p=0.396). **B.** Female mutant and WT were studied at similar ages (WT n=7, Mut n=8, p=0.290). **C.** Male mutant mice weighed significantly less than WTs (WT n=6, Mut n=7 p=0.047). **D.** Female mutant mice weighed significantly less than female WTs (WT n=7, Mut n=8, p=0.001). E. Weights of bladders drained of urine were similar in WT and Mut males (WT n=6, Mut n=7, p=0.262). **F.** Female Mut bladders were significantly heavier than WTs (WT n=7, Mut n=8, p=0.001). In all figures, WT males are represented by black plain squares, Mut males by purples open squares, WT females by black plain circles and Mut females with purple open circles. Results are expressed as mean±SEM. WT compared with Mut *p<0.05, ***p<0.001.

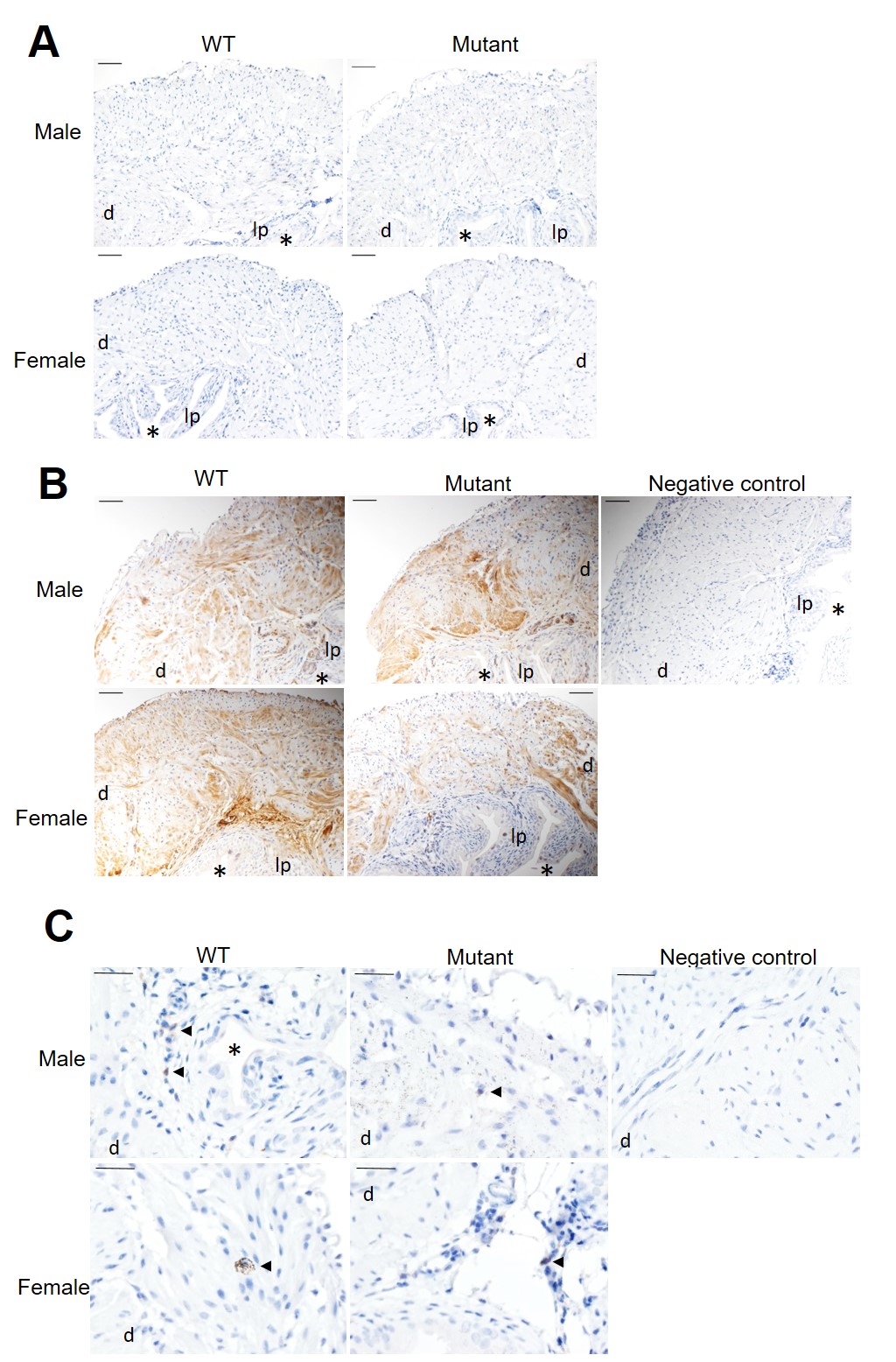

**Figure S2. Histology of WT and *Lrig2* Mut male and female bladders. A.** Representative images of bladder dome sections after staining with haematoxylin. **B**. Representative images of bladder sections after immunostaining for TGFβ1. **C.** Representative images of bladder dome stained for the macrophage marker F4/80. Macrophages are indicted by the arrowheads. The ‘negative controls’ have the primary antibody omitted. ‘d’ indicates detrusor, lp indicates lamina propria * indicates lumen.Scale bars are 200 µm in A and B and 100 µm in C.

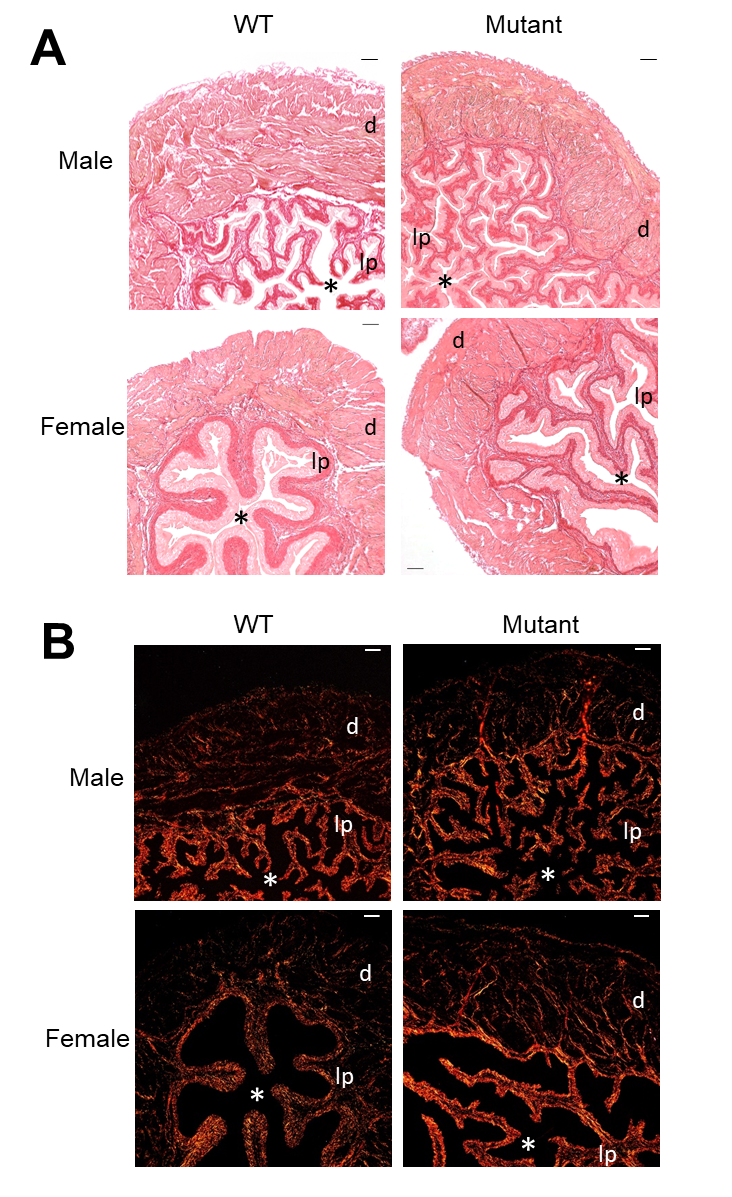

**Figure S3. PSR staining of bladder dome sections. A.** Representative pictures of bladder section after PSR staining and imaging with brighfield microscopy. **B.** Representative pictures of bladder section after PSR staining and imaging with polarised light. d indicates the detrusor, lp indicates the lamina propria and the asterisk indicates the lumen. Scale bar 200 μm.

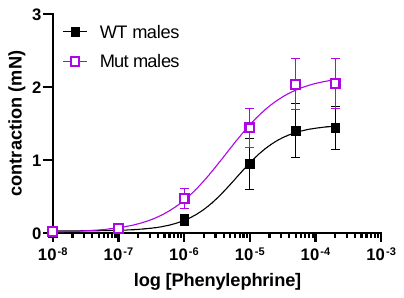

**Figure S4. Male outflow responses to phenylephrine.** Concentration-dependence of the contraction response to PE in WT (n=6) and Mut (n=6) male outflows. Points and bars represent mean ± sem. Curves are the best fits to the Hill equation with EC_50_= 6.23 µM and Emax = 1.5 mN for WT and EC_50_= 4.34 µM and Emax = 2.1 mN for Mut outflows. A 2-Way ANOVA with repeated measures did not show differences between WT and Mut.
